# Testing behaviour may bias observational studies of vaccine effectiveness

**DOI:** 10.1101/2022.01.17.22269450

**Authors:** Paul Glasziou, Kirsten McCaffery, Erin Cvejic, Carys Batcup, Julie Ayre, Kristen Pickles, Carissa Bonner

## Abstract

**Backgroun:** Recent observational studies have suggested that vaccines for the omicron variant of SARS-Cov2 may have little or no effect in preventing infection. However, the observed effects may be confounded by patient factors and preventive behaviours or vaccine-related differences in testing behaviour. To assess the potential degree of confounding, we aimed to estimate differences in testing behaviour between unvaccinated and vaccinated populations.

**Methods:** We recruited 1,526 Australian adults for an online randomised study about COVID testing between October and November 2021, and collected self-reported vaccination status and three measures of COVID-19 testing behaviour. We examined the association between testing intentions and vaccination status in the cross-sectional baseline data of this trial.

**Results:** Of the 1,526 participants (mean age 31 years): 22% had a COVID-19 test in the past month and 61% ever; 17% were unvaccinated, 11% were partially vaccinated (1 dose), 71% were fully vaccinated (2+ doses). Fully vaccinated participants were twice as likely (RR 2.2; 95% CI 1.8 to 2.8) to report positive COVID testing intentions than those who were unvaccinated (p<.001). Partially vaccinated participants had less positive intentions than those fully vaccinated (p<.001) but higher intentions than those who were unvaccinated (p=.002).

**Discussion:** For all three measures vaccination predicted greater COVID testing intentions. If the unvaccinated tested at half the rate of the vaccinated, a true vaccine effectiveness of 30% could appear to be a “negative” observed vaccine effectiveness of -40%. Assessing vaccine effectiveness should use methods to account for differential testing behaviours. Test negative designs are currently the preferred option, but its assumptions should be more thoroughly examined.

## Introduction

Recent observational studies have suggested that vaccines for the omicron variant of SARS-Cov2 may have little or no effect in preventing infection while being effective in preventing hospitalisation. For example, a Danish study^1^ found low vaccine efficacy against omicron infections: 55% for the BNT162b2 and 37% for the mRNA-1273 vaccine. However, for longer follow up (after 90 days) the “effectiveness” observed was negative (less than 0), triggering reports than vaccines paradoxically might increase omicron infections, which was picked up by multiple blogs and subsequently fact-checked by AP News^2^. However, differences in testing behaviour may bias such observational studies of vaccines effectiveness.

As for all observational studies of vaccines, the observed effects may be confounded by other patient factors and preventive behaviours such as social distancing and mask wearing^3^. However, vaccine-related differences in COVID-19 testing behaviour may also cause differences in case detection (though this is less likely to impact hospitalisations). For example, if the unvaccinated were half as likely as the vaccinated to get tested for respiratory symptoms, they would also be half as likely to be detected as COVID-19 cases. This differential testing would artefactually dilute estimates of vaccine effectiveness (or could even create a spurious negative effect from vaccines, as seen in the Danish study). The “test negative” design,^4^ - which stratifies for health seeking behaviour by patients who test negative as controls - is a common method to reduce the bias from health care-seeking behaviour differences between vaccinated and unvaccinated people, but relies on assumptions about severity and test accuracy^5^. To assess the degree of confounding potentially caused by any differential testing behaviour, we aimed to assess any differences in past and intended testing behaviour between unvaccinated and vaccinated populations.

## Methods

We recruited 1,526 Australian adults (430 men, 1064 women, 32 non-binary or not reported) for an online randomised study about COVID testing between October and November 2021 (www.anzctr.org.au/Trial/Registration/TrialReview.aspx?id=382318) using methods we have detailed previously^6^. We collected self-reported vaccination status (unvaccinated, 1 dose or 2 doses) and three measures of COVID-19 testing behaviour. We considered two vaccine doses as ‘fully vaccinated’ at the time of assessment as third dose boosters were only available for high-risk workers (e.g. health care workers) in Australia at this timepoint. We examined the association between testing intentions and vaccination status across the sample with Pearson chi-squared tests, and non-parametric linear-by-linear tests for trend. Analyses were conducted using Stata/BE v17.0.

## Results

The 1,526 participants (mean age 31 years) were from all Australian states: 22% had a COVID-19 test in the past month and 61% ever; 17% were unvaccinated, 11% were partially vaccinated (1 dose), 71% were fully vaccinated (2+ doses). Fully vaccinated participants were twice as likely (RR 2.2; 95% CI 1.8 to 2.8) to report positive COVID testing intentions than those who were unvaccinated (p<.001) – Table. Partially vaccinated participants had less positive intentions than those fully vaccinated (p<.001) but higher intentions than those who were unvaccinated (p=.002).

**Table.**
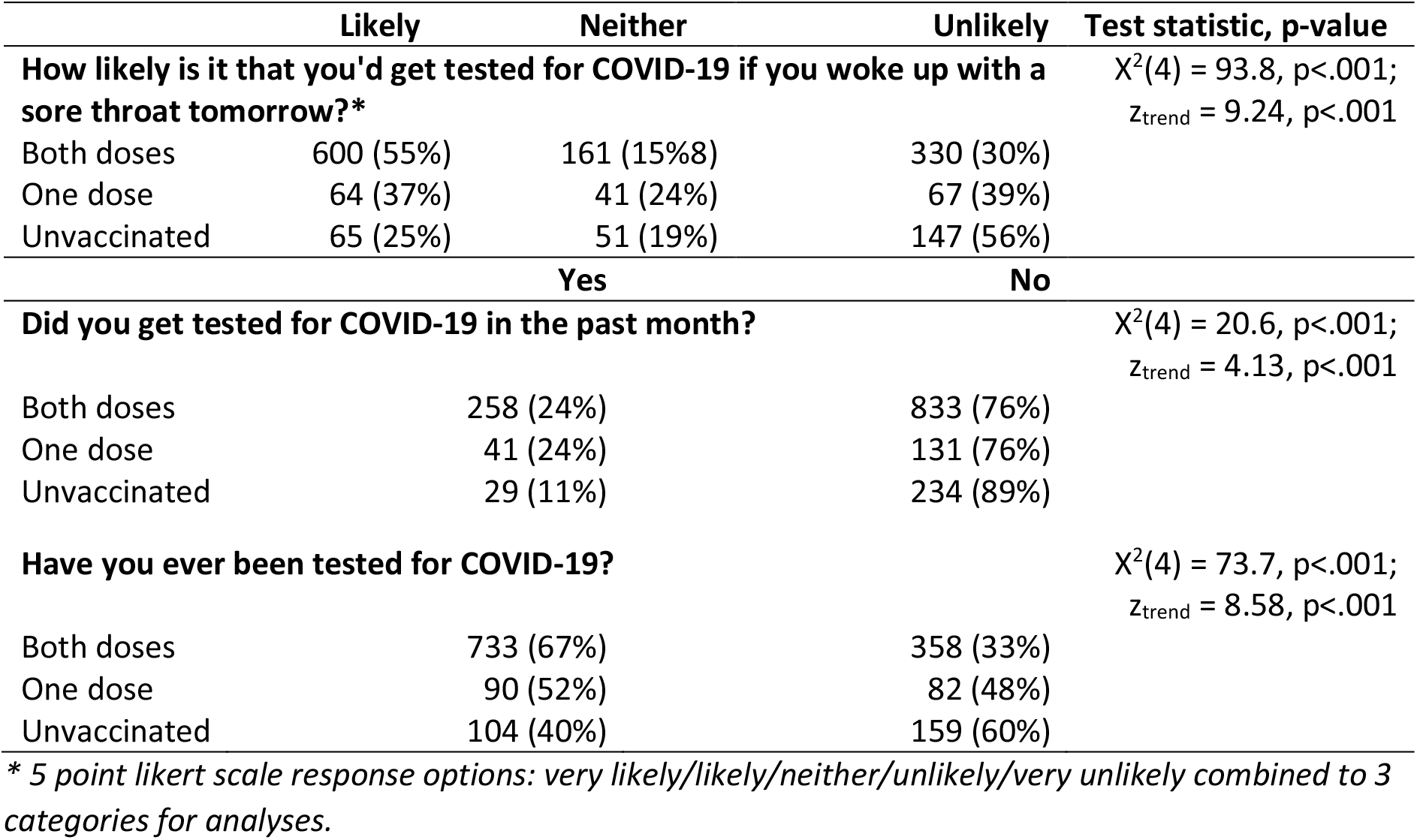
Association between vaccination status and COVID testing (i) intentions and (ii) self reported behaviours (n=1,526). Data are shown as frequencies (row percentages).

Fully vaccinated participants were also twice as likely (RR 2.1; 95% CI 1.5 to 3.0) to report being tested in the past month than those who were unvaccinated (p<.001).

## Discussion

For all three measures (intention, self reported testing in past month or ever) vaccination predicted greater COVID testing intentions and behaviours in the vaccinated compared to the unvaccinated. If confirmed in other studies, this has behavioural difference has both policy and research methods implications.

Methods for observational studies of influenza vaccine effectiveness adjust for several important confounders, but the spectrum of confounding behaviours in the current SARS-CoV2 pandemic may be different, particularly substantial differential testing behaviour. If unvaccinated people were detected as cases at half the rate of the vaccinated (as seen for the “tested in the past month” in Table), then a true vaccine effectiveness of 30% would lead to a “negative” observed vaccine effectiveness of -40% (as seen in the Danish study for Moderna vaccine at 91-150 days). A recent report from the Statens Serum Institut in Denmark^7^ also noted lower testing rates of the unvaccinated compared to the vaccinated of a similar magnitude to that in our study.

A recent WHO technical brief^8^ of vaccine effectiveness against the omicron variant did include the Danish study but the majority of the studies were “test negative” designs. For example, the UK HSA analysis^9^ using a test negative design estimated that the vaccine effectiveness against symptomatic disease was 26%, 2 to 18% (for 2-24 weeks versus 25+ weeks post-dose), and 63% for 1, 2, and 3 doses of vaccine respectively. However, for hospitalisation the vaccine effectiveness was substantially better at 52%, 52 to 72%, and 88% respectively. However, such test negative studies may still be biased by test seeking behaviour if there are differences in severity between cases and non-cases^5^.

Assessing vaccine effectiveness against SARS-CoV2 infection should use methods to account for differential testing behaviours. Test negative designs are currently the preferred option, but its assumptions should be more thoroughly examined.

## Data Availability

Please for information about the data

## Data Availability

Please for information about the data

## Acknowledgements and Funding

This study was not specifically funded, but in-kind support was provided by authors with research fellowships. PG is supported by a National Health and Medical Research Council (NHMRC) Investigator Grant (#1080042).CB is supported by a National Health and Medical Research Council (NHMRC)/Heart Foundation Early Career Fellowship (#1122788). RD is supported by a University of Sydney fellowship (#197589). KM is supported by a National Health and Medical Research Council (NHMRC) Principal Research Fellowship (#1121110).

## Notes

### Competing Interest Statement

The authors have declared no competing interest.

### Author Declarations

University of Sydney Human Ethics, approval number # 2020/781

